# SARS-CoV-2 transmission, vaccination rate and the fate of resistant strains

**DOI:** 10.1101/2021.02.08.21251383

**Authors:** Simon A. Rella, Yuliya A. Kulikova, Emmanouil T. Dermitzakis, Fyodor A. Kondrashov

## Abstract

Vaccines are thought to be the best available solution for controlling the ongoing SARS-CoV-2 pandemic^1,2^. However, the emergence of vaccine-resistant strains^3–6^ may come too rapidly for current vaccine developments to alleviate the health, economic and social consequences of the pandemic^7,8^. To quantify and characterize the risk of such a scenario, we created a SIR-derived model^9,10^ with initial stochastic dynamics of the vaccine-resistant strain to study the probability of its emergence and establishment. Using parameters realistically resembling SARS-CoV-2 transmission, we model a wave-like pattern of the pandemic and consider the impact of the rate of vaccination and the strength of non-pharmaceutical intervention measures on the probability of emergence of a resistant strain. We found a counterintuitive result that the highest probability for the establishment of the resistant strain comes at a time of reduced non-pharmaceutical intervention measures when most individuals of the population have been vaccinated. Consequently, we show that a period of transmission reduction close to the end of the vaccination campaign can substantially reduce the probability of resistant strain establishment. Our results suggest that policymakers and individuals should consider maintaining non-pharmaceutical interventions^7,11,12^ throughout the entire vaccination period.

## Main

Vaccines are among the most effective public health measures against infectious disease^1^. Their track record brings hope that SARS-CoV-2 may soon be under control^13^ as a consequence of a plethora of vaccine development efforts^14–17^. A potential cause of concern is the low rate of vaccine production and administration^18^ coupled with reports of new strains with higher transmission rates^3,4,19^ and even with potential for some degree of vaccine resistance^5,6,8,20^. A number of models considered the dynamics of the spread of a vaccine-resistant strain in the population^9,10,21,22^. However, to our knowledge, the interplay of the population vaccination rate with the stochastic dynamics of emergence of a resistant strain has been discussed^23^, but not formally modeled. Specifically, a concern is whether a combination of vaccination and transmission rates can create positive selection pressure on the emergence and establishment of resistant strains^24,25^. To address this issue, we implemented a model to simulate the probability of emergence of a resistant strain as a function of vaccination rates and changes in the rate of virus transmission, resembling those caused by non-pharmaceutical interventions and behavioural changes. We then performed a number of simulations based on realistic parameters to study the likelihood and pattern of the emergence of a resistant strain. Finally, we considered possible countermeasures to reduce the probability of the establishment of the resistant strain in the population.

We implemented a modification of a SIR model^10,26^ that included additional states to study the interplay of the rate of vaccination, rate of transmission and the likelihood of emergence of resistant strains (**Fig. 1a**). In addition to other states, individuals could be vaccinated (V), infected by the resistant strain (I_r_), or simultaneously be vaccinated and infected with the resistant strain (I_r_^V^). In the model, susceptible individuals (S) are infected by the wildtype strain at a rate of β and infected individuals recover at a rate of γ or die at a rate of δ. At each time step, a fraction θ of all non-infected individuals is vaccinated and with some probability *p*, an infected individual becomes infected with a resistant strain. Overall, our model included 8 character states and 6 transition parameters between them (**Fig. 1a, Extended Data Table 1**).

**Fig. 1.**
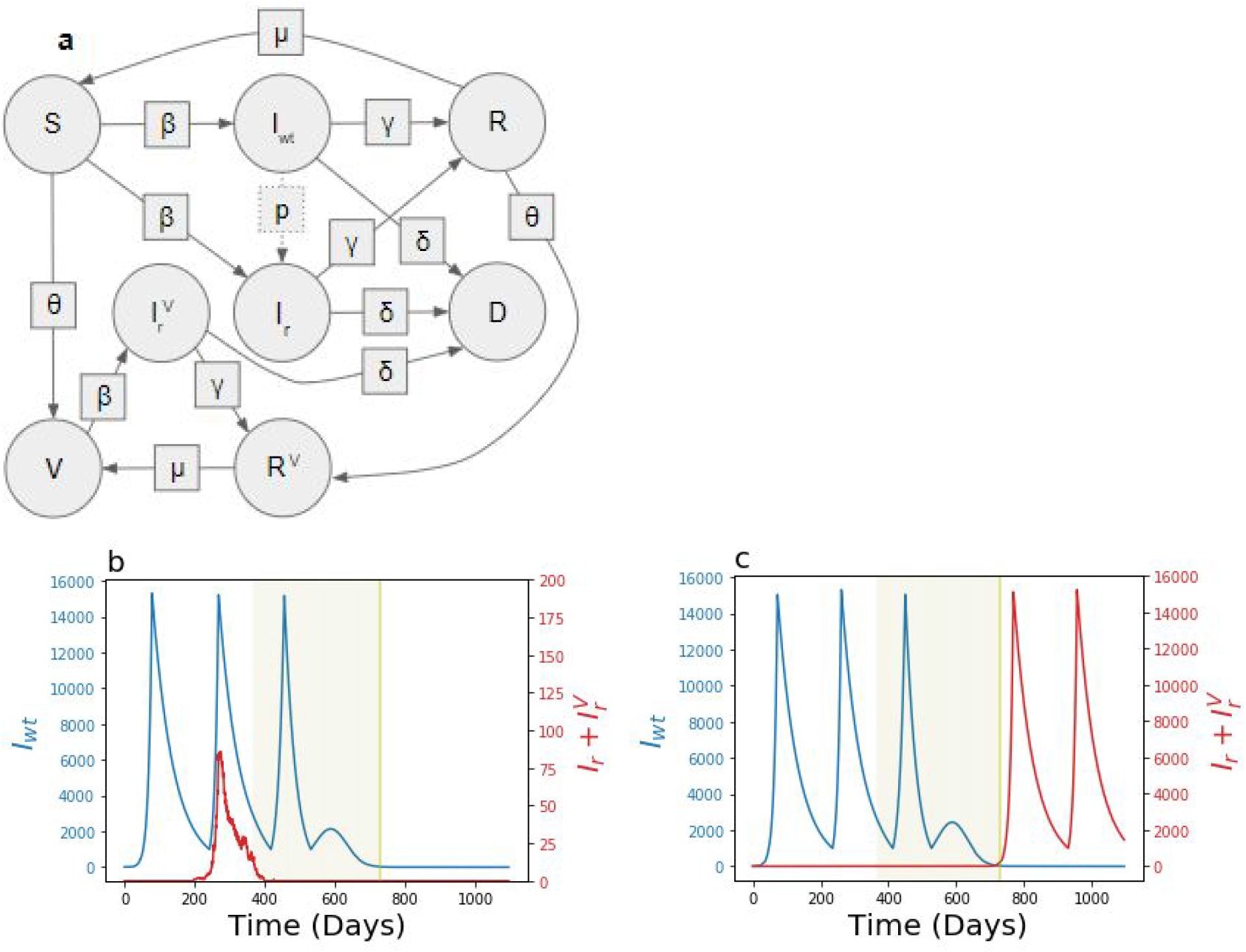
The states, transition parameters and dynamics. **a**, States are shown in circles and transition parameters in squares. The transition parameter, μ, is the rate at which individuals lose natural immunity and *p*, is the probability that an individual infected with the wildtype strain transmits a resistant strain, so it is not a deterministic parameter. Example dynamics of the number of individuals infected with the wildtype (blue) and resistant strains (red) for p = 10^−6^, θ_0_ = 1/365 and F = 15000. The period of vaccination is highlighted (green). Under the same parameters the resistant strain may emerge and go extinct, **b**, or become established, **c**.

The rate of transmission in the course of a pandemic is typically cyclical^27–29^ due to government interventions^29,30^, behavioural changes^31–33^, and environmental^34,35^ and other factors^36,37^. Generally, the number of infected individuals is wave-like, guided by periods of high rate of transmission, followed by periods of a low rate of transmission^27,28,38,39^. We thus varied β_t_, the rate of virus transmission to reflect this cyclical behavior (**Figs. 1b**,**c**). A high rate of transmission (β_h_ = 0.18, equivalent to the effective reproduction number of R_h_ = 2.52) was alternated with a low rate (β_l_ = 0.055 or R_l_ = 0.77), which broadly reflected the observed rates of transmission in various countries affected by the SARS-CoV-19 pandemic with and without lockdown measures, respectively^39–41^. The low rate of transmission was triggered in the model when the number of individuals infected with any strain reached F = (I_wt_ +I_r_ +I_r_^V^). Transition from a low rate of transmission back to a high rate occurred at a constant value of (I_wt_ +I_r_ +I_r_^V^) = 1000. We also considered a mode when the transition from low to high rate of transmission occurred at (I_wt_ +I_r_ +I_r_^V^) = F/2, however, the results were not qualitatively different and are not reported here.

SIR-like models frequently consider only deterministic dynamics^10^. However, the emergence of a new strain is an inherently stochastic process under extensive influence of genetic drift^42,43^. Therefore, we incorporated a stochastic stage into our model to allow for genetic drift in the early phases of population dynamics of the resistant strains. The growth rate of the number of individuals infected with the wildtype strain at time *t* was determined deterministically by (β*S/N - γ - δ)I_wt_. By contrast, when the frequency of the resistant strain in the population is low, the number of transmissions of the resistant strain was drawn from a Poisson distribution with a mean of (β*I_r_*(S+V)/N)dt [44]. However, when the frequency of the resistant strain is greater than 1000 individuals (0.01% of the population), making it highly unlikely to disappear by stochastic forces, the dynamics are treated deterministically in the same manner as the wildtype strain.

The initial stages of the vaccine-resistant strain propagation consisted of emergence, the appearance of the first individual with the infected strain, establishment, the time point when the number of infected individuals reached 1000, and extinction, when the number of resistant strain infected individuals returned to zero. The impact of three parameters on the resistant strain propagation were explored, the probability of the emergence of the resistant strain (p), the speed of vaccination (θ) and the initiation of periods of lower rate of transmission (F). All other parameters were constants and their values were chosen to be broadly reflecting a realistic set of parameters that approximate the available data for the SARS-CoV-2 pandemic, μ =1/180, γ = 0.99*1/14 and δ = 0.01*1/14 (see **Extended Data Table 1**). The model was run to simulate a population of 10,000,000 individuals over three years with the vaccination starting after the first year. Higher values of *p* had a predictably^45^ positive effect on the probability of the establishment of the resistant strain (**Extended Data Figs. 1-3**), but depended on the rate of vaccination (θ) and low transmission initiation (F) in a complex manner (**Fig. 2, Extended Data Figs. 1-3**). In a specific parameter range the probability of establishment of the resistant strain was increased by a factor of two (**Figs. 2d**,**e**).

**Fig. 2.**
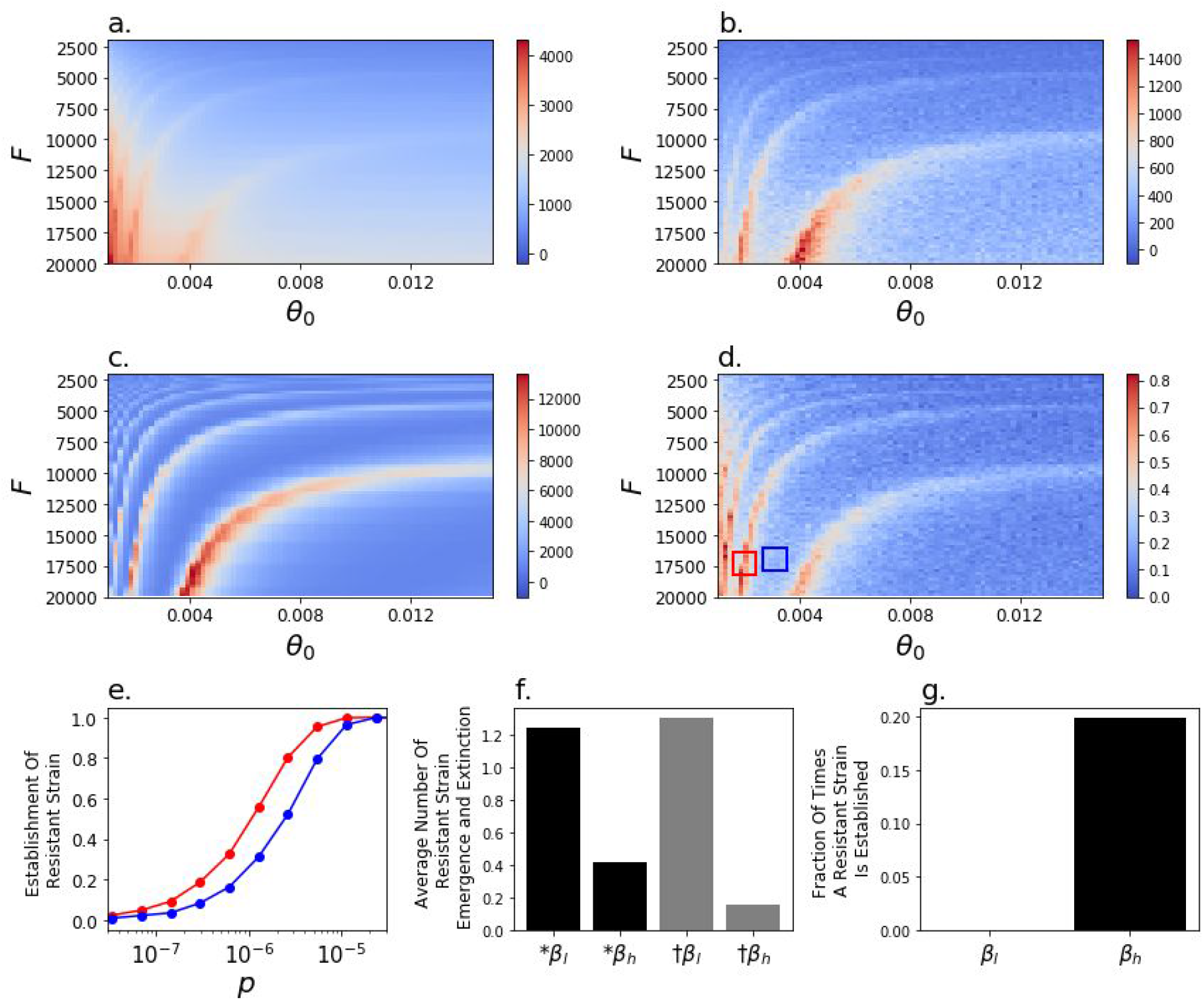
Impact of the rate of vaccination and initiation of low rate of transmission on model dynamics. The cumulative death rate from the **a**, wildtype and **b**, resistant strains, **c**, the number of wildtype-strain infected individuals at t_v60_, the point in time when 60% of the population is vaccinated and **d**, the probability of resistant strain establishment, for p=10^−6^. **e**, The probability of emergence of the resistant strain as a function of the probability of emergence, p, in the parameter ranges of θ and F indicated in corresponding colour boxes in **d. f**, The average number of times of 8×10^6^ simulation runs during which a resistant strain emerges (black) or goes extinct (grey) during periods of low (β_l_) or high (β_h_) transmission for p = 10^−6^. **g**, A resistant strain was never observed to establish during periods of low transmission (β_l_) for p = 10^−6^.

The behaviour of the emergence, establishment and extinction of the resistant strain in the population bears striking resemblance to the population genetics problem addressing the survival of a beneficial allele in a growing population^46,47^. To understand the stochastic dynamics of the resistant strain in the model, it is therefore instructive to consider the underlying mechanism in population genetics terms. Unless the rate of mutation is zero, or infinitesimally low, new variants will emerge in the population at a rate of p*I_wt_. When the rates of transmission of the wildtype and the resistant strain are equal, the probability that the resistant strain will go extinct, 1 - Q_t_, can be approximated by

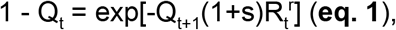

where s is the selection advantage of the resistant strain^42,48^ and R_t_^r^ = (S+V)β_t_/N(γ + δ). Therefore, even when there is no selective advantage of the resistant strain (s = 0) over the wildtype strain but the rate of transmission is high (R_t_^r^ > 1), the likelihood that a new mutation is lost from the population is small (∼10% for β_h_ = 0.18, or R_0_ = 2.52). By contrast, when the rate of transmission is low (R_t_^r^ < 1, which is the case during the low transmission periods in the model), the probability of extinction of the resistant strain by genetic drift is substantial^42,43^. The results of our model are consistent with theory, such that the resistant strain emerges during periods of high and low transmission rates, but goes extinct with higher probability during periods of low transmission (**Fig. 2f**). Furthermore, under the parameters of our model the resistant strain becomes established only during periods of high transmission (**Fig. 2f**).

The complex influence of the speed of vaccination, θ, and initiation of low transmission period, F, on the dynamics of establishment of the resistant strain (**Figs. 2d**) may, therefore, be driven by the overlap of the vaccination period and the periodicity of the cycles of the number of infected individuals driven by F (**Fig, 1b**.**c**). The coincidence of a high number of vaccinated individuals and a high rate of transmission has two effects on the resistant strain. First, as mentioned previously, because the rate of transmission is high, the emerging resistant strain is not lost through genetic drift (see also^46,49^). Second, a high number of vaccinations creates a selective advantage of the resistant strain over the wildtype strain^25^. The effective reproductive number of the wildtype versus the resistant strains, R_t_^wt^/R_t_^r^, is (V+S)/S, which is the selective advantage 1+s in **eq. 1**. Thus, when V is large the resistant strain has a growth advantage over the wildtype strain, contributing to its establishment in the population towards the end of the vaccination campaign. Taken together, the highest probability for establishment of the resistant strain for a given *p* is reached when V, I_wt_ and β_t_ (and the corresponding R_t_^r^) are large (**Fig. 2c, eq. 1**).

Indeed, when p = 10^−6^, in those cases when the resistant strain becomes established, its initial time of emergence frequently occurs at around the time when 60% of the population is vaccinated (**Fig. 3**). Therefore, we then tested the influence of a single intervention triggering at a single extraordinary period of low transmission centred around 60% of vaccinated individuals in the population (**Fig. 3**). We varied the duration of this intervention, T, ranging from one week to 120 days and considered three rates of transmission, β_l_ = 0.055 (R_0_ = 0.77), 0.03 (R_0_ = 0.42) and 0.01 (R_0_ = 0.14). Both parameters decrease probability of establishment of the resistant strain with the length of the intervention having a relatively stronger effect (**Fig. 3. Extended Data Figs. 1-7**).

**Fig. 3.**
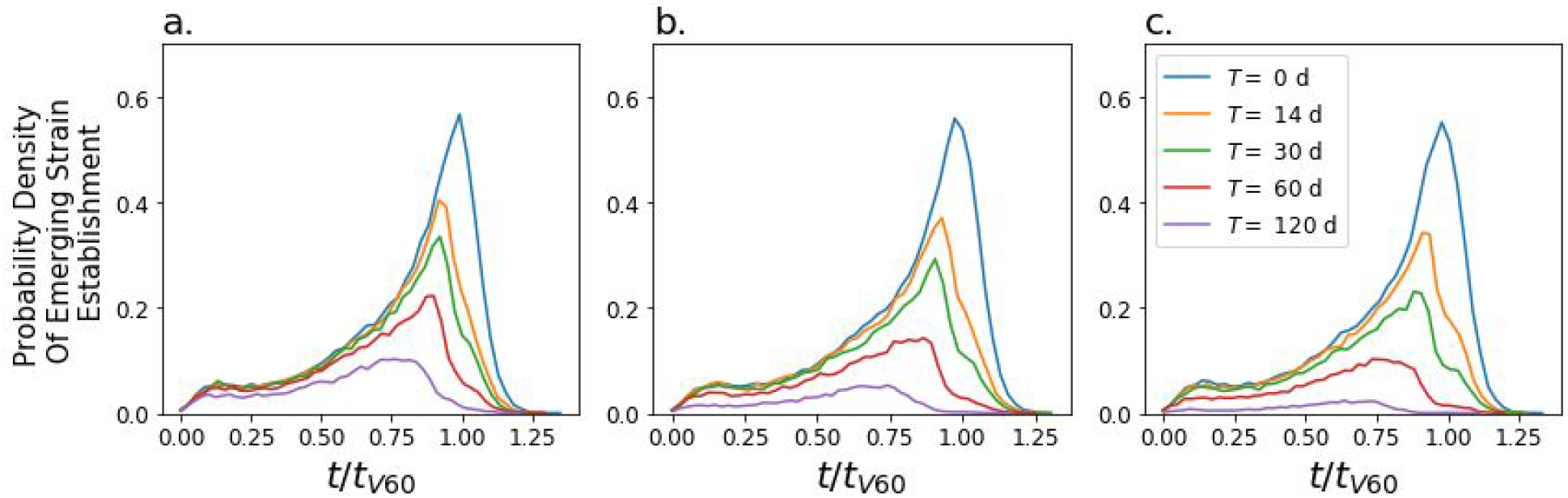
Time of initial emergence of a resistant strain that has become established. Probability density that the resistant strain emerges as a function of time since the start of the simulation, *t*, rescaled by the time at which 60% of the individuals are vaccinated, *t*_v60_, averaged across simulations with θ (0.001 through 0.015), F (2,000 through 20,000) and p = 10^−6^. Without any extraordinary periods of low transmission (blue line) the peak of the likelihood of emergence of a new strain is at *t*/*t*_V60_ = 1. The likelihood of emergence of a resistant strain can be reduced by an extraordinary period of low transmission centered at *t*/*t*_v60_ = 1 with a stronger reduction when such period is longer, T (colour-coded), or when the rate of transmission is more strongly reduced **a**, β_l_ = 0.055, **b**, β_l_ = 0.03, **c**, β_l_ = 0.01.

In conclusion, our model suggests three specific risk factors that favour the emergence and establishment of a vaccine-resistant strain that are intuitively obvious: high probability of initial emergence of the resistant strain, high number of infected individuals^50^ and low rate of vaccination^51^. By contrast, a counterintuitive result of our analysis is that the highest risk of resistant strain establishment occurs when a large fraction of the population has already been vaccinated but the transmission is not controlled (see ref. [52] for empirical data consistent with this result in influenza). Indeed, it seems likely that when a large fraction of the population is vaccinated, especially the high-risk fraction of the population (aged individuals and those with specific underlying conditions) policy makers and individuals will be driven to return to pre-pandemic guidelines and behaviours conducive to a high rate of virus transmission. However, the establishment of a resistant strain at that time may lead to serial rounds of resistant strain evolution with vaccine development playing catch up in the evolutionary arms race against novel strains.

The results of our model provide several qualitative implications for the strategy forward in the months of vaccination. In our model, the probability of emergence of a resistant strain in one individual per day was in the range of 10^−5^ to 10^−8^ for a population of 10^7^ individuals. For the entire human population of ∼10^10^ that probability would be 10^−8^ to 10^−11^, which does not seem improbably large. As of February 2021, ∼10^9^ individuals have been infected by SARS-CoV-2 [53] with an average 14 days of sickness per individual^54^, so >10^10^ number of total days of infected individuals. Furthermore, highly mutated strains may emerge as a result of long shedding in immunocompromised individuals, a rare but realistic scenario^55–57^. Taken together, the emergence of a partially or fully vaccine-resistant strain and its eventual establishment appears inevitable. However, as vaccination needs to be ahead of the spread of such strains in similar ways to influenza^25^, it is necessary to reduce the probability of establishment by a targeted effort to reduce the virus transmission rate towards the end of the vaccination period before the current vaccines become ineffective. Conversely, lack of non-pharmaceutical interventions at that time can increase the probability of establishment of vaccine-resistant strains. For example, plans to vaccinate individuals with a high risk of a fatal disease outcome followed by a drive to reach herd immunity while in uncontrolled transmission among the rest of the population is likely to greatly increase the probability that a resistant strain is established, annulling the initial vaccination effort. Another potential risk factor may be the reversion of vaccinated individuals to pre-pandemic behaviours that can drive the initial spread of the resistant strain.

One simple specific recommendation is to keep transmission low even when a large fraction of the population has been vaccinated by implementing acute non-pharmaceutical interventions (i.e. a strict lockdown) for a reasonable period of time, to allow emergent lineages of resistant strains to go extinct through stochastic genetic drift. Additional factors that may make these measures even more effective are: (i) increased and widespread testing, (ii) rigorous contact tracing, (iii) high rate of viral sequencing of positive cases^52,58^ and (iv) travel restrictions. Finally, while our model formally considers only one homogenous population, our data also suggest that delays in vaccination in some countries relative to others will make the global emergence of a vaccine-resistant strain more likely. Thus, a truly global vaccination effort may be necessary to reduce the chances of a global spread of a resistant strain.

## Data Availability

The results of our modeling are reported in the manuscript in full and the primary code is available on Github

https://github.com/Simon-Re/mut-vacc

## Acknowledgements

We thank Alexey Kondrashov, Nick Machnik, Raimundo Julian Saona Urmeneta, Gasper Tkacik and Nick Barton for fruitful discussions. The opinions expressed in this document are exclusively of the authors and, therefore, do not necessarily coincide with those of the Banco de España or the Eurosystem. ETM is supported by the Swiss National Science and Louis Jeantet Foundation. The work of FAK was in part supported by the ERC Consolidator grant (771209—CharFL).

## Author Contributions

ETM and FAK formulated the question. SAR and YAK formulated the model. SAR programmed and ran the model. All authors jointly analyzed the results and wrote the manuscript.

## Code availability

https://github.com/Simon-Re/mut-vacc

## Extended Data Figures and Legends

**Extended Data Fig. 1.**
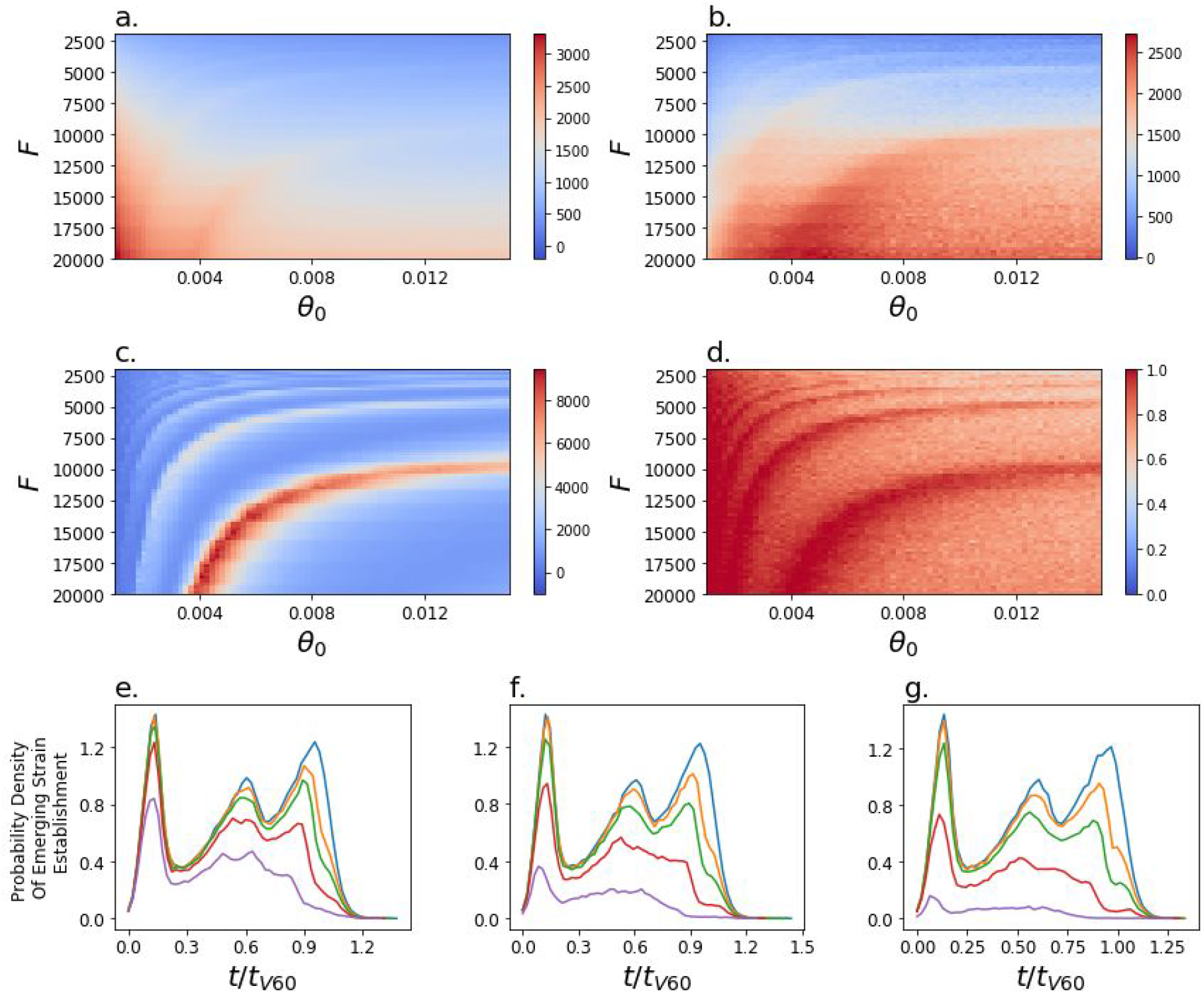
Impact of the rate of vaccination and initiation of low rate of transmission on model dynamics for p = 10^−5^. The cumulative death rate from the **a**, wildtype and **b**, resistant strains, **c**, the number of wildtype-strain infected individuals at t_v60_, the point in time when 60% of the population is vaccinated and **d**, the probability of resistant strain establishment. **e-g**, Probability density that the resistant strain emerges as a function of time since the start of the simulation, *t*, rescaled by the time at which 60% of the individuals are vaccinated, *t*_v60_, summed across simulations with θ (0.001 through 0.015), F (2,000 through 20,000). The impact of the extraordinary low transmission period centered at *t*/*t*_v60_ = 1 on the likelihood of emergence of the resistant strain as a function of the duration of that period, T (colour-coded), and the intensity of the reduction of transmission **e**, β_l_ = 0.055, **f**, β_l_ = 0.03, **g**, β_l_ = 0.01.

**Extended Data Fig. 2.**
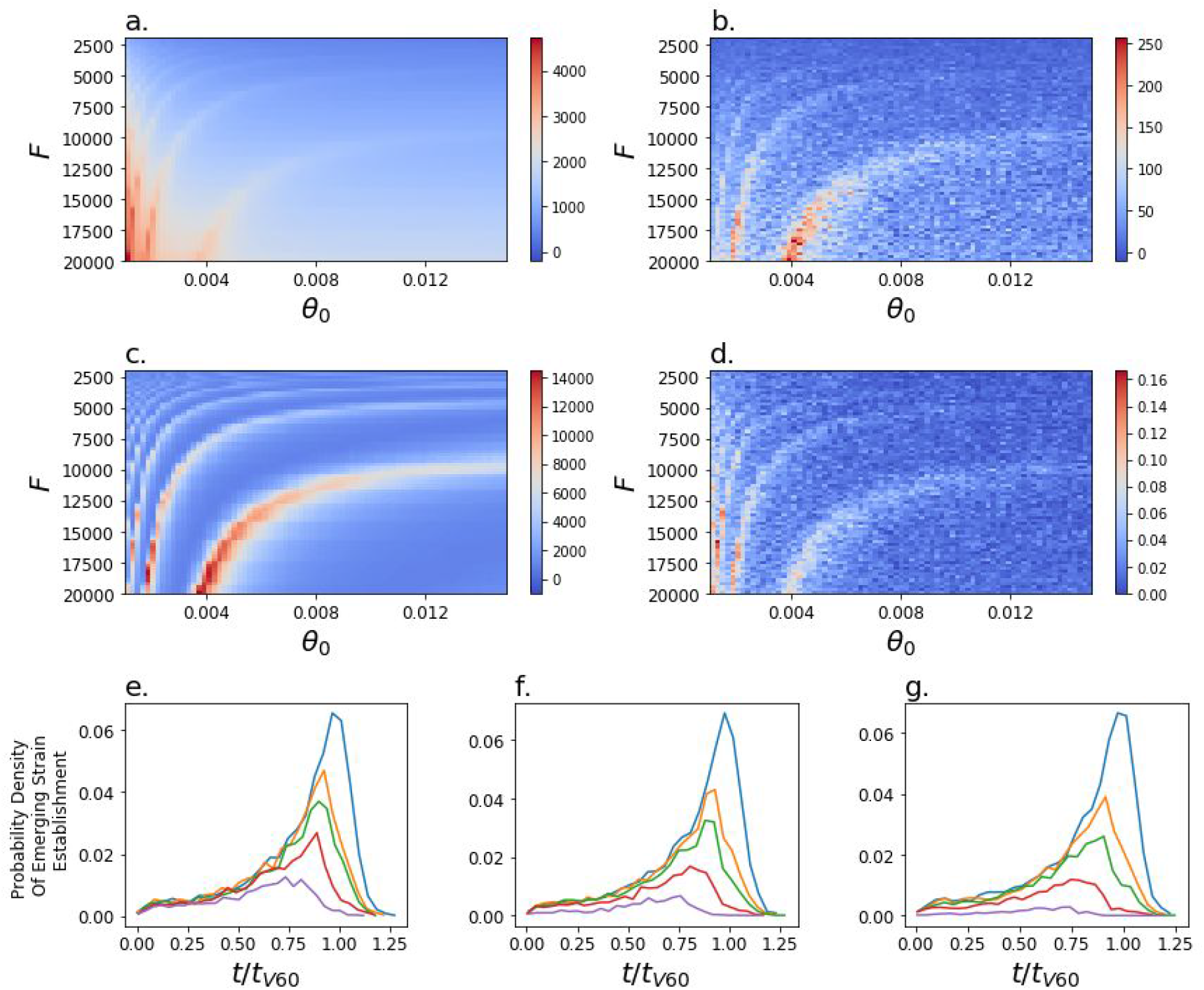
Impact of the rate of vaccination and initiation of low rate of transmission on model dynamics for p = 10^−7^. The cumulative death rate from the **a**, wildtype and **b**, resistant strains, **c**, the number of wildtype-strain infected individuals at t_v60_, the point in time when 60% of the population is vaccinated and **d**, the probability of resistant strain establishment. **e-g**, Probability density that the resistant strain emerges as a function of time since the start of the simulation, *t*, rescaled by the time at which 60% of the individuals are vaccinated, *t*_v60_, summed across simulations with θ (0.001 through 0.015), F (2,000 through 20,000). The impact of the extraordinary low transmission period centered at *t*/*t*_v60_ = 1 on the likelihood of emergence of the resistant strain as a function of the duration of that period, T (colour-coded), and the intensity of the reduction of transmission **e**, β_l_ = 0.055, **f**, β_l_ = 0.03, **g**, β_l_ = 0.01.

**Extended Data Fig. 3.**
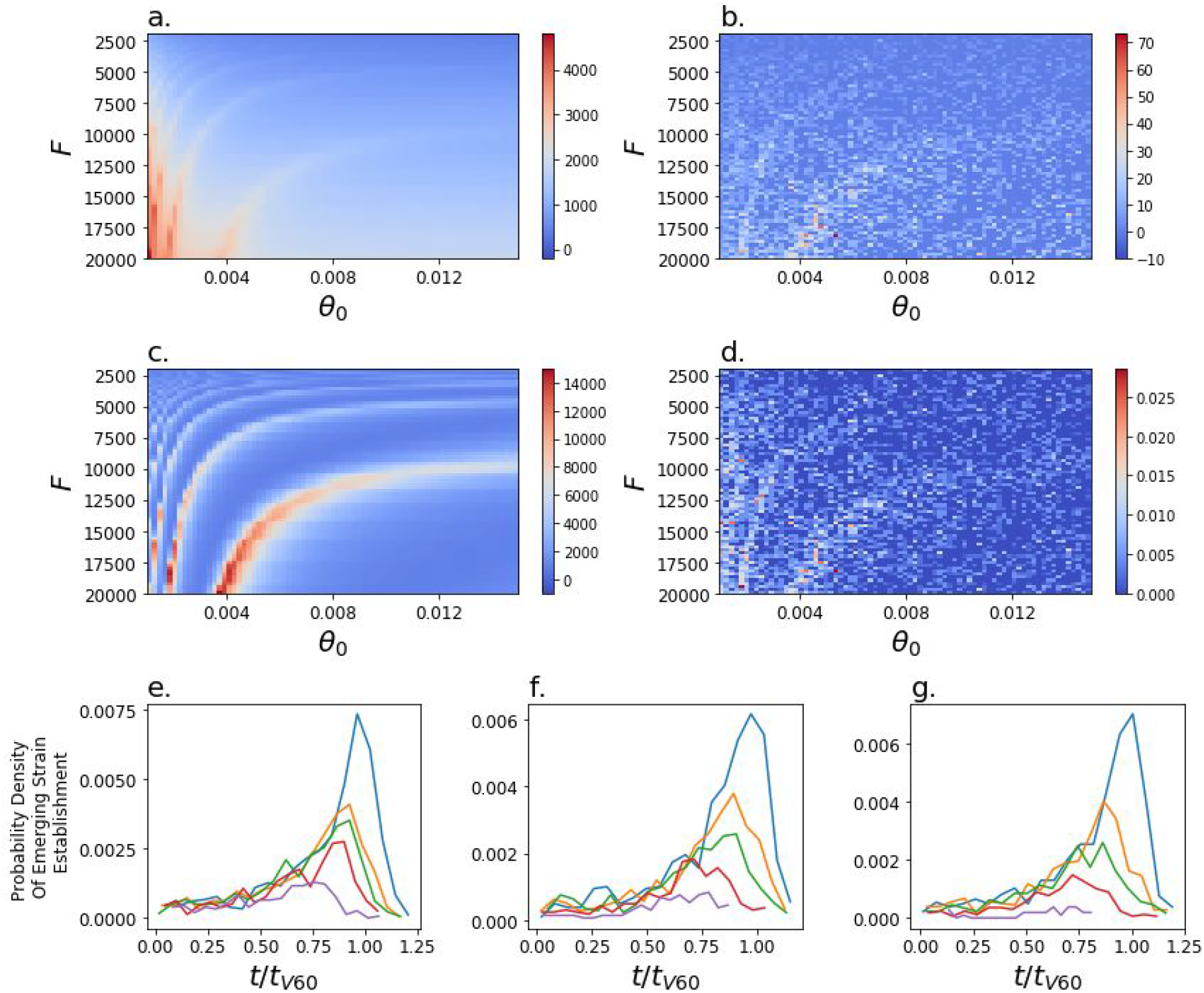
Impact of the rate of vaccination and initiation of low rate of transmission on model dynamics for p = 10^−8^. The cumulative death rate from the **a**, wildtype and **b**, resistant strains, **c**, the number of wildtype-strain infected individuals at t_v60_, the point in time when 60% of the population is vaccinated and **d**, the probability of resistant strain establishment. **e-g**, Probability density that the resistant strain emerges as a function of time since the start of the simulation, *t*, rescaled by the time at which 60% of the individuals are vaccinated, *t*_v60_, summed across simulations with θ (0.001 through 0.015), F (2,000 through 20,000). The impact of the extraordinary low transmission period centered at *t*/*t*_v60_ = 1 on the likelihood of emergence of the resistant strain as a function of the duration of that period, T (colour-coded), and the intensity of the reduction of transmission **e**, β_l_ = 0.055, **f**, β_l_ = 0.03, **g**, β_l_ = 0.01.

**Extended Data Fig. 4.**
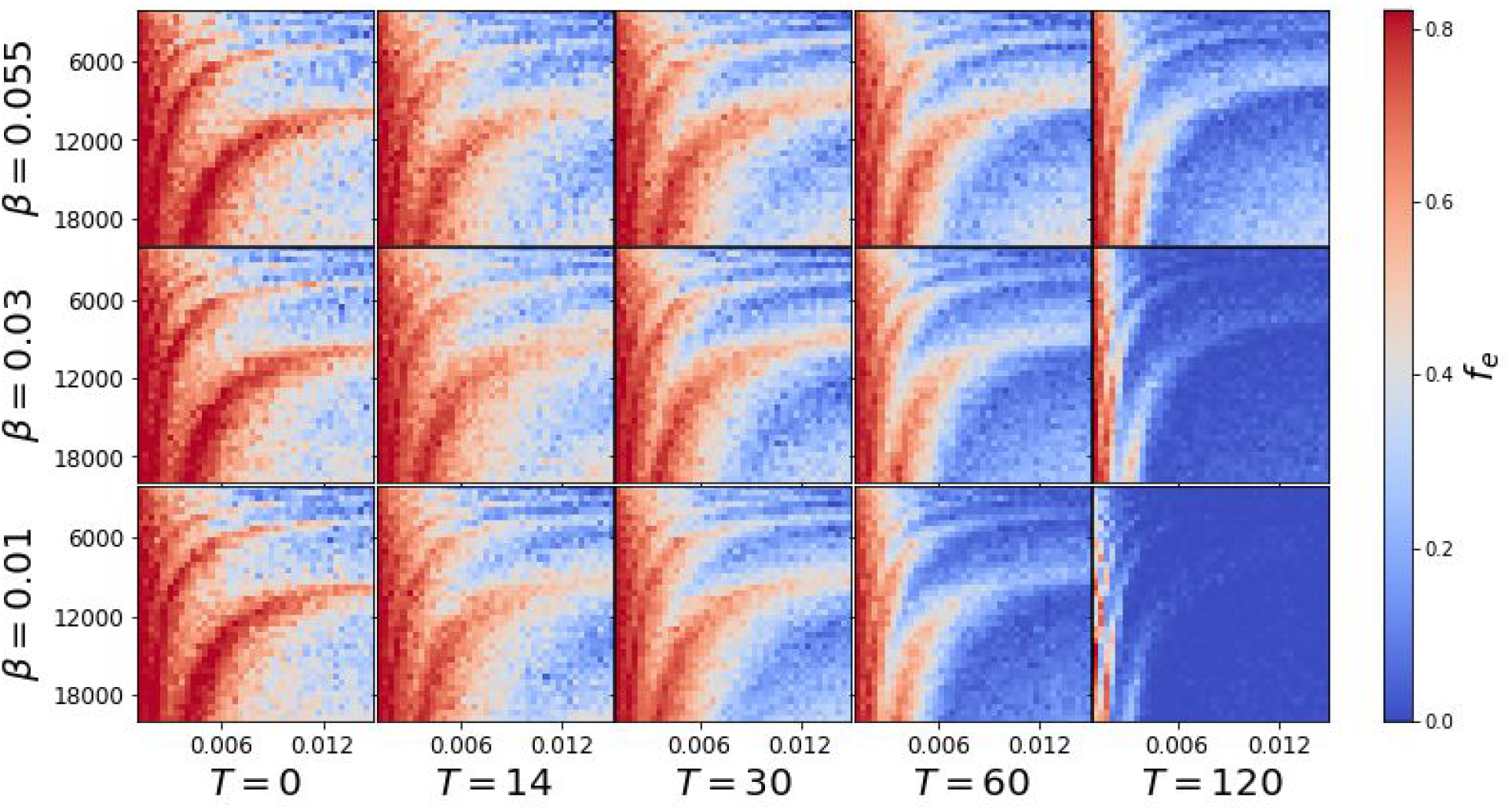
The probability of establishment of the resistant strain for p = 10^−5^. The influence of low transmission period centered at *t*/*t*_v60_ = 1 on probability of establishment of the resistant strain as a function of the duration of that period, T, and the intensity of the reduction of transmission, β.

**Extended Data Fig. 5.**
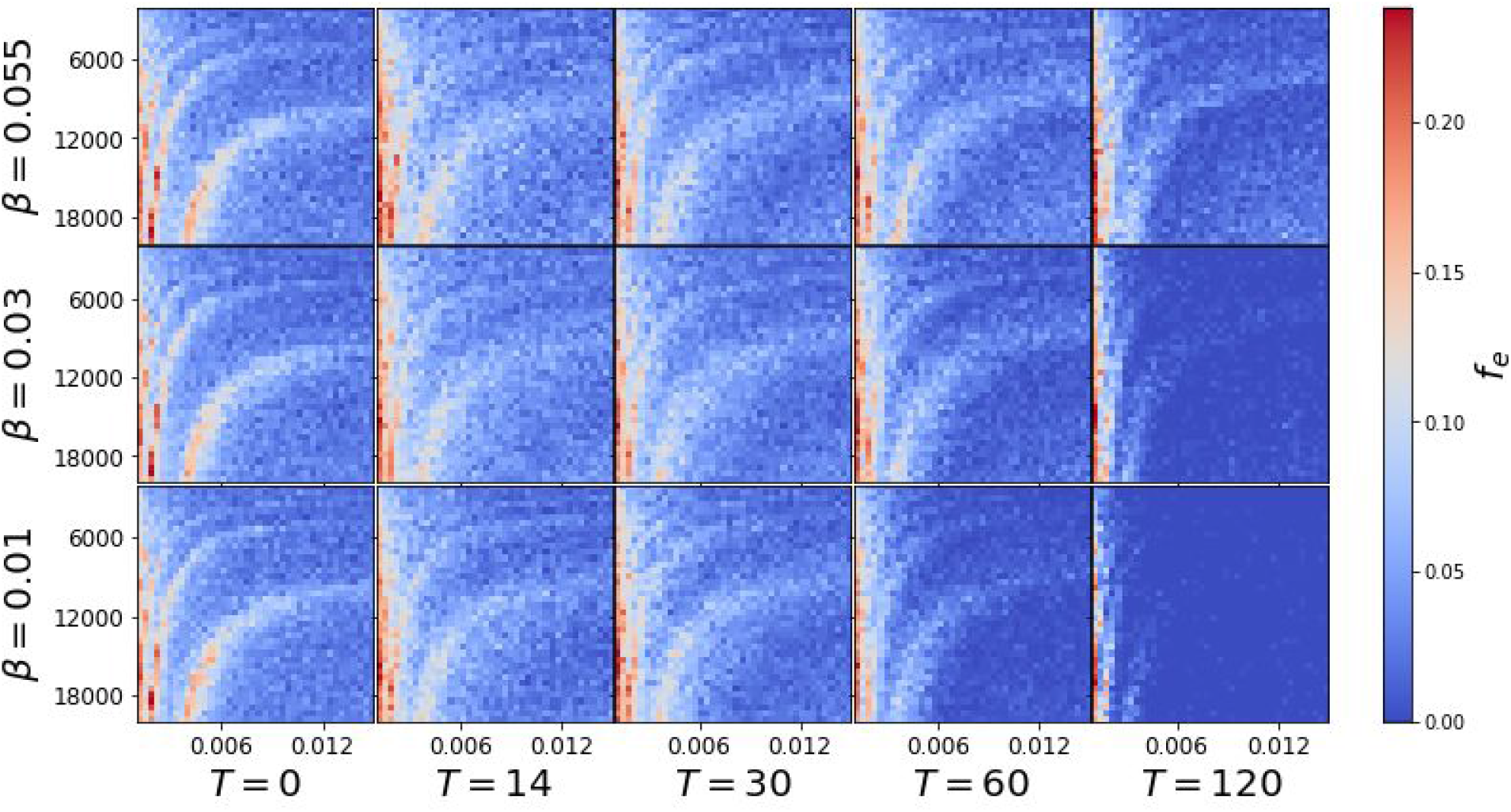
The probability of establishment of the resistant strain for p = 10^−6^. The influence of low transmission period centered at *t*/*t*_v60_ = 1 on probability of establishment of the resistant strain as a function of the duration of that period, T, and the intensity of the reduction of transmission, β.

**Extended Data Fig. 6.**
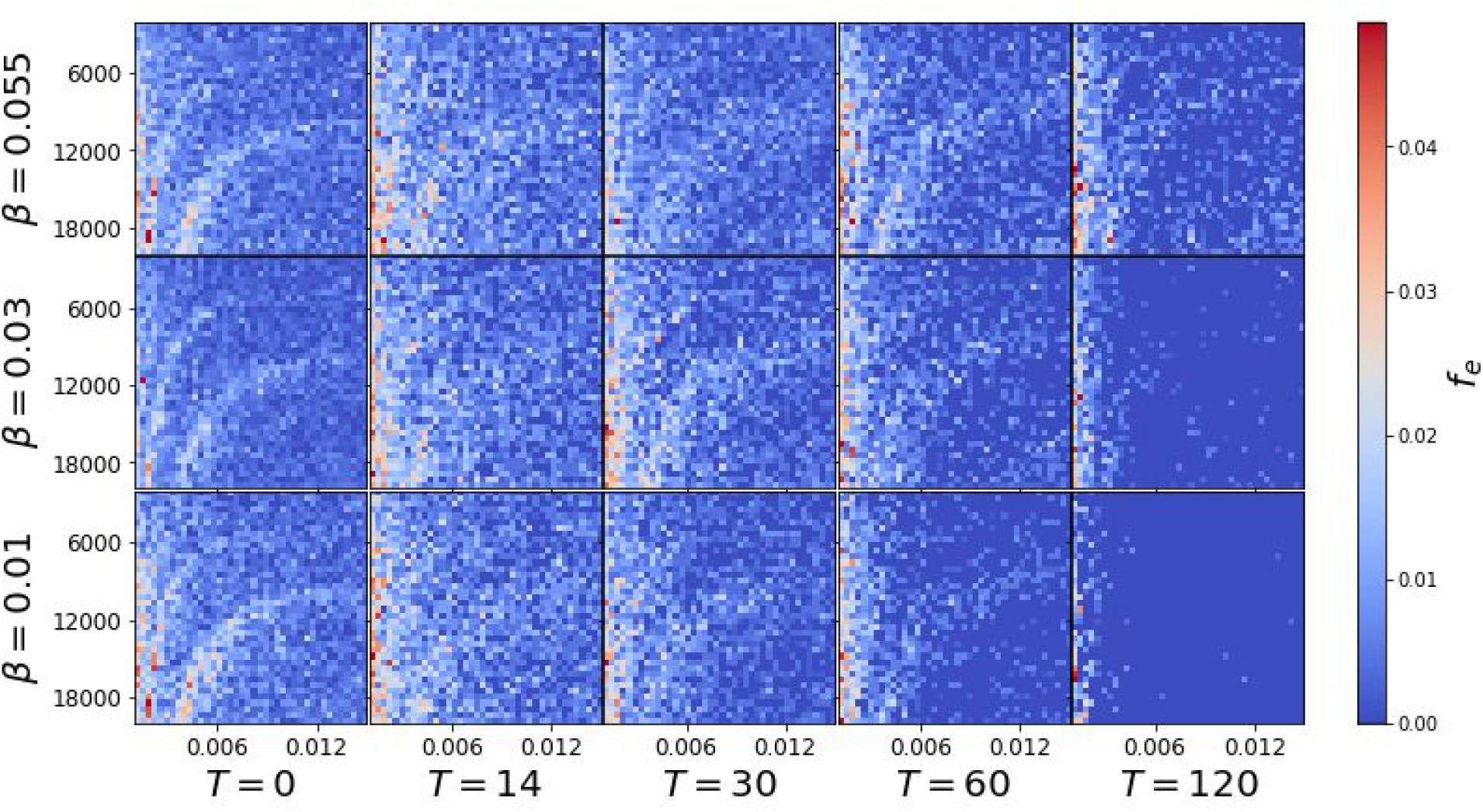
The probability of establishment of the resistant strain for p = 10^−7^. The influence of low transmission period centered at *t*/*t*_v60_ = 1 on probability of establishment of the resistant strain as a function of the duration of that period, T, and the intensity of the reduction of transmission, β.

**Extended Data Fig. 7.**
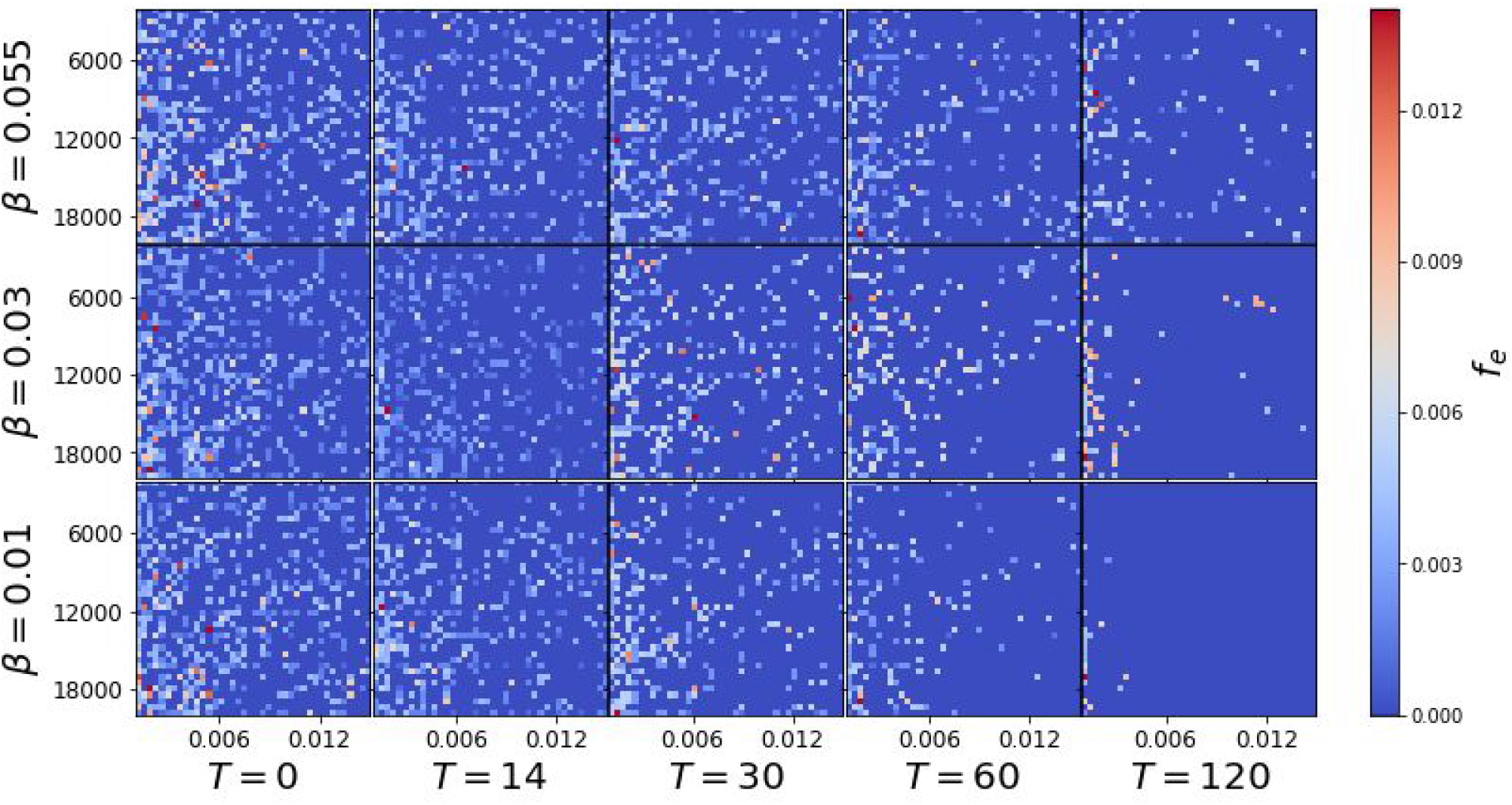
The probability of establishment of the resistant strain for p = 10^−8^. The influence of low transmission period centered at *t*/*t*_v60_ = 1 on probability of establishment of the resistant strain as a function of the duration of that period, T, and the intensity of the reduction of transmission, β.

## Methods

Our extension of the SIR Model features 8 distinct states. Susceptible, *S*, and recovered, *R*, individuals are vaccinated over time to become vaccinated, *V*, or recovered vaccinated, *R*^*V*^. Susceptible individuals can become infected with the wildtype, *I*_*wt*_, or the resistant virus strain, *I*_*r*_. While the vaccinated population is immune against the wildtype, it can be a target to the resistant strain, 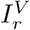. After a while any infected individual recovers or dies, *D*. Finally, we assume that the recovered population retains natural immunity towards both strains, but becomes susceptible again with some small rate, *µ*. In our model, immunity against the wildtype strain gained through vaccination is not lost during the entire model period of 3 years.

The total number of individuals, *N*, in the population remains constant at 10,000,000.

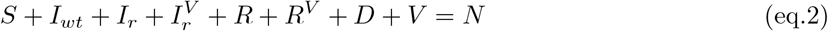

In the limit of large population sizes, the full dynamics without mutations can be described by the following set of differential equations.

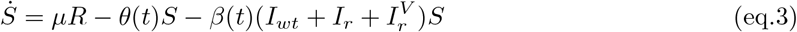

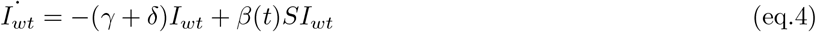

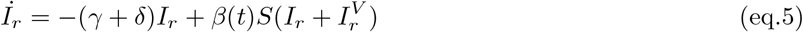

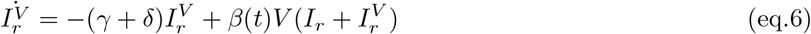

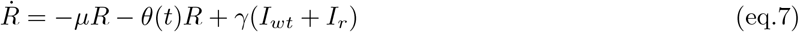

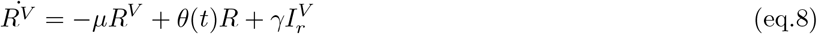

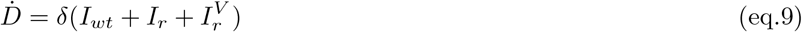

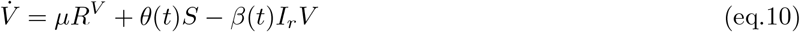

Or 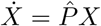, with 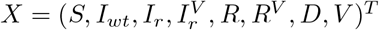

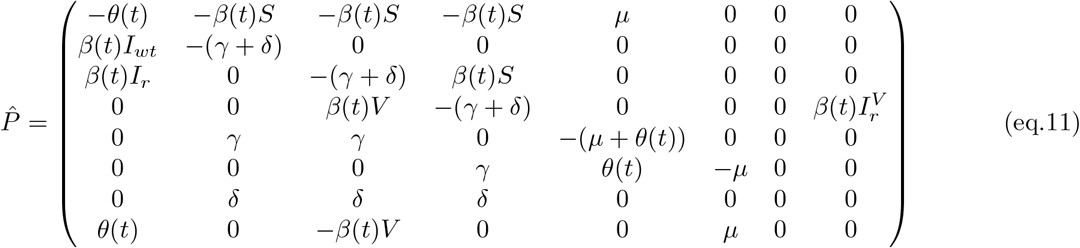

The dynamics are influenced by the following constant parameters: the recovery rate, *γ*, the death rate, *δ*, and the rate at which natural immunity is lost, *µ*. Additionally we introduce a time dependent transmission rate *β*(*t*) and a function *θ*(*t*), which controls the speed of vaccination.

### Time Dependent Transmission Rate

*β*(*t*) switches between high and low transmission rates^27,28,38,39^. The model begins with a period of a high rate of transmission, *β*_*h*_*/N*. A low transmission rate, *β*_*l*_*/N*, is initiated when the fraction of individuals infected with any strain, 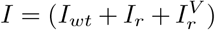, reaches the value of *F*. Transition from a period of low to high transmission occurs when *I* = 1000.

### Vaccination

Vaccination is modelled as a mostly linear function with saturation. *h* denotes the number of individuals in the population that are never vaccinated. A maximum of *N* − *h* individuals can be vaccinated at the end of the vaccination program. The constant *k* controls the saturation of the vaccination speed once the number of susceptible individuals is significantly depleted. The state dependent vaccination speed *θ*(*t*) is given as:

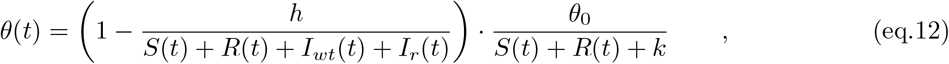

where *θ*_0_ can take different values and *h* and *k* are chosen to be small (see Extended Data Table 1).

### Integration Method

The deterministic differential equations eq.11 were numerically solved using an Euler Forward Integration Scheme, with time step Δ*t*, measured in days.

### Resistant Strain

Each day and for every *I*_*wt*_ individual, there is a small probability *p*, that a vaccine-resistant strain emerges in that individual. Then this individual switches from state *I*_*wt*_ to state *I*_*r*_. Conversely, any *I*_*r*_ individual can revert back to the wildtype strain population *I*_*wt*_ with the same probability *p*. Each time step the number of mutations is drawn from a Poisson distribution with mean Δ*tpI*_*wt*_, for mutations to the resistant strain, or Δ*tpI*_*r*_, for mutations to the wildtype strain.

### Stochastic and Deterministic Regimes

The population dynamics of a rare variant is an inherently stochastic process^42,43^. We can formally treat the spread of a disease in our model as a stochastic birth-death process. In the following we illustrate this with the number of wildtype infections *I*_*wt*_ as an example. In each infinitesimally small time step *dt*, there is a probability *βSI*_*wt*_*dt*, that the wildtype population *I*_*wt*_ grows by 1, *I*_*wt*_ → *I*_*wt*_ + 1, while the susceptible population is decreased, *S* → *S* − 1. Similarly, with probability (*γ* + *δ*)*I*_*wt*_*dt, I*_*wt*_ is reduced by 1, *I*_*wt*_ → *I*_*wt*_ − 1, while the number of recovered or dead grows by 1. We carefully model small populations, *I*_*wt*_ < *N* ^*^, using a stochastic Tau-Leaping Algorithm^44^. We choose a fixed time step size *τ*, that is equal to the time step of the Euler Integrator Δ*t*. For very small *I*_*wt*_ Tau Leaping Algorithm can produce negative population^59^. This stems from the fact, that the number of events *K* that occur in time *τ* is drawn from a Poisson Distribution, that always assigns a non-zero probability for any *K* > *I*_*wt*_. We reduce the chances of such a scenario, by solving the exact SSA Gillespie Algorithm for very small *I*_*wt*_ that is below a critical size *N*_*c*_^44^.

For large *I*_*wt*_, larger than some *N* ^*^, this stochastic process can by approximated with the limiting differential eq.4 and an Euler Integration Scheme. Once *I*_*wt*_ ≥ *N* ^*^, we consider that the resistant strain of the virus is established in the population and we continue modelling it using the deterministic equation^10^.

### Parallel Evaluation Of Deterministic And Stochastic Variables

In our model we can then evaluate deterministic and the stochastic dynamics in parallel. While small populations of infected individuals are treated as stochastic, other variables, such as the number of susceptible individuals, *S*, are evaluated within the deterministic regime. While the infection numbers of the wildtype or the emergent strain are in the stochastic regime (< *N* ^*^), the corresponding terms that contain the wildtype infections *I*_*wt*_ or the emergent infections 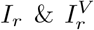 are removed from the deterministic rate equations. When *I*_*wt*_ or 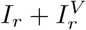 grows above the threshold value *N* ^*^, the corresponding population of infected individuals is treated as deterministic.

#### Sources of Errors

Finally, we discuss some sources of errors in our simulation: (1) Depending on the time step Δ*t* the Euler Integration Scheme is not exact. In most of our simulations, we choose a time step of one day, Δ*t* = 1*d*. (2) Using the deterministic rate equations for the infection numbers in eq.11 is an approximation to the exact stochastic dynamics given by a birth-death model. The quality of this approximation is given by the threshold value *N* ^*^. In our model we use a threshold of *N* ^*^ = 1000. (3) When *I*_*wt*_ or 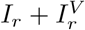 trespasses the threshold *N* ^*^ from above, the populations of infected individuals changes from being treated as a real number (the mean field average) to being treated as a natural number. We truncate the mean field average with a floor function and treat the remainder as part of the recovered population, *I*_*wt*_ → ⌊*I*_*wt*_*⌋* & *R* → *R* + *I*_*wt*_ ⌊*I*_*wt*_ ⌋. (4) The Tau Leaping algorithm is an approximation to the exact SSA Gillespie Algorithm, that allows faster evaluation with a constant step size *τ*. Increasing the threshold value *N*_*c*_ increases the accuracy of the model (Supplementary Figure 1). (5) As discussed above, it can rarely happen that a population of infected individuals drops below 0 in one leap. If this happens we disregard the draw and redraw from the same Poisson distribution. (6) Finally, while the time step of the deterministic model Δ*t* and *τ* are chosen to be equal, for population sizes above *N*_*c*_, the SSA algorithm acts on exponentially distributed waiting times *τ*_*SSA*_ between reactions^44^. This introduces errors, if *τ*_*SSA*_ ≫ Δ*t*, because the interaction rates may change, while we wait for the next reaction to occur.

In order to determine a range of acceptable values of *N*_*c*_, we ran our simulation for a period of *T* = 200 days, initially loading the system with *I*_*wt*_ = 200 wildtype carriers. For multiple values of *N*_*c*_ and no mutations, we compared the results of disease survival with the analytical solution derived for the birth death process,

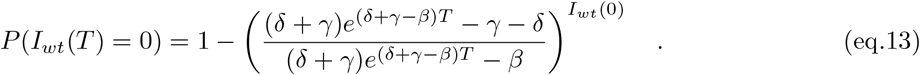

We pick *N*_*c*_ = 100 days, which gives us a reasonably small error.

**Supplementary Figure 1:**
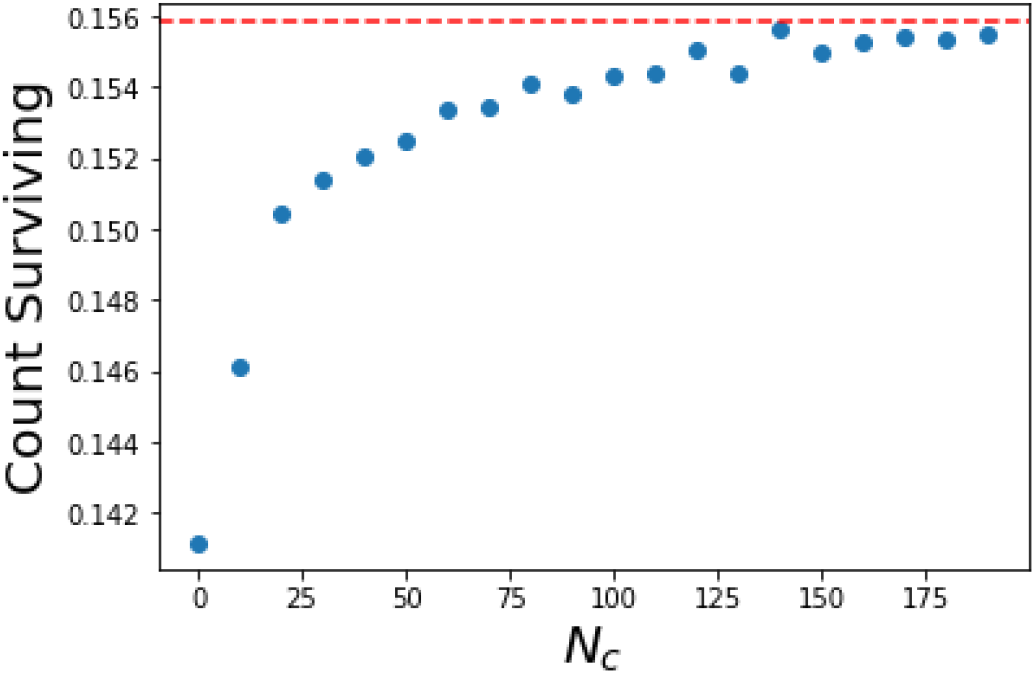
Shown is the fraction of surviving strains after *T* = 200 days in 10^7^ runs, first initialized with *I*_*wt*_ = 200 infected individuals. The red dashed line shows the expected fraction of surviving strains, as computed with eq.13. The stochastic algorithm becomes exact, if no Tau Leaping is employed and instead the whole simulation is evaluated using the Gillespie SSA scheme.

## Assumptions and Choice of Parameters

The model is run for a total time of three years, with vaccination starting one year into the model. We assume that the wildtype and emergent strains have the same infectivity (*β* is the same for both strains). We assume that infection by any one strain provides immunity to both, reflecting that many vaccines carry only the Spike protein of the SARS-CoV-2 virus and it may be easier to escape immunity provided by the vaccine than the immunity provided by infection. We also assume that the immune response provided by the vaccine is more permanent and that immunity provided by infection, *µ*, decays in 0.5 years^13,60^ after recovery. Both of these assumptions influence the model when the number of infected individuals becomes large, which is unlikely for realistic average rates of transmission across the simulated time.

We assume that susceptible and recovered individuals have an equal chance to be vaccinated, *θ*_0_. We also assume that the infection-recovery rate, *γ*, and infection-fatality rate, *δ* are the same for the wildtype and mutated strains.

We regulate the rates of transmission exogenously in the model, with the rate of transmission (*β*) switching between a high rate, *β*_*h*_ and a low rate, *β*_*l*_, when the total number of individuals infected with either strain reaches a threshold parameter. This threshold parameter, *F*, simultaneously reflects the impact of all non-pharmaceutical interventions and behavioural changes of individuals. We operate under the assumption that vaccine efficacy not only impacts disease manifestation but also blocks transmission at the same rate, which is a reasonable assumption based on previous vaccine performance but has not yet been demonstrated.

In Extended Data Table 1 we present the choice of parameters for the model, the ones that were constant and those that were varied, including their boundaries.

**Extended Data Table 1:** Model Parameters

| Parameter | Value | Comments |
| --- | --- | --- |
| Fixed parameters |  |  |
| Population size, $N$ | 10,000,000 individuals | |
| Cut-off for stochastic mode, $N^*$ | 1000 | |
| Cut-off for Gillespie algorithm, $N_c$ | 100 | |
| Recovery rate, $\gamma$ | $0.99 \times 1/14$ | Average disease duration 14 days<sup>54</sup> |
| Death rate, $\delta$ | $0.01 \times 1/14$ | Infection-fatality rate is 1%<sup>61,62,63</sup> |
| Loss of immunity rate, $\mu$ | $1/180$ | On average in 180 days<sup>64,65</sup> |
| Share of non-vaccinated, $h$ | 0.01 | 1% of population |
| Saturation parameter, $k$ | 0.01 | |
| Transmission rate, $\beta = \{\beta_l, \beta_h\}$ | $\{0.055, 0.18\}$ | $R_0 = \{0.77, 2.52\}$ <sup>39,40,41</sup> |
| Bound for initiation of high transmission | 1,000 individuals |  |
| Varying parameters |  |  |
| Bound for initiation of low transmission, $F$ | 2,000 to 20,000 individuals | |
| Probability of emergence of resistant strain, $p$ | $1e-8$ to $1e-5$ | Daily for every infected |
| Vaccination speed, $\theta_0$ | 0.001 to 0.015 | 0.1% to 1.5% daily |

